# Probiotic supplements reduce antipsychotic-induced metabolic disturbances in drug-naïve first-episode schizophrenia

**DOI:** 10.1101/2021.02.16.21251872

**Authors:** Dongyu Kang, Fengyu Zhang, Ye Yang, Chenchen Liu, Jingmei Xiao, Yujun Long, Jing Huang, Xingjie Peng, Weiyan Wang, Xiaoyi Wang, John M. Davis, Jingping Zhao, Renrong Wu

## Abstract

Probiotic supplements have demonstrated efficacy in improving metabolic abnormalities and may prevent antipsychotic-induced metabolic disturbance and weight gain. A few studies in rodents have found that antipsychotic-induced metabolic dysfunctions are associated with the altered composition of gut microbiota. Here, we conducted a randomized-controlled clinical trial to determine the effectiveness and safety of probiotic supplements on antipsychotic-induced metabolic disturbance and weight gain. Patients with drug-naïve first-episode schizophrenia were randomized to receive either olanzapine plus probiotics or olanzapine monotherapy and scheduled to evaluate with follow-ups for clinical and metabolic profiles. After a treatment of 12 weeks with addition of probiotics, the increase of mean fasting insulin was significantly lower than olanzapine monotherapy. Insulin resistance increased considerably in the olanzapine plus probiotics, but also significantly lower than olanzapine monotherapy. We noted a difference in the increase in body mass index and body weight between treatments at a nominal level of significance, but it became non-significant after adjusting for appetite increase. Probiotics concurrently used with olanzapine is effective and safe in attenuating antipsychotic-induced elevation of fasting insulin and insulin resistance, but not the weight gain in drug-naïve first-episode schizophrenia. Further study is warranted to assess the longer-term maintenance of efficacy and safety.

## 1. Introduction

The second-generation antipsychotics (SGAs) are widely used as first-line medications to treat and manage patients with schizophrenia. However, antipsychotics-induced weight gain and metabolic dysregulations such as hyperglycemia and hyperlipidemia are of significant concern (Newcomer, 2007). These side effects not only affect the patients’ compliance with antipsychotic medications but also cause substantial medical morbidities such as diabetes, cardiovascular disease, stroke, and premature death (De Hert et al., 2011; Foley and Morley, 2011). Antipsychotic drugs have sharply increased in the prescription of children and adolescents and are shown to cause significant weight gain and risk for type 2 diabetes (Maayan and Correll, 2011). Of the SGAs, olanzapine is one of the commonly used and has better efficacy than other drugs in patients with chronic schizophrenia (Lieberman et al., 2005). In a large cohort study about the SGAs Treatment Indications, Effectiveness, and Tolerability in Youth Study (SATIETY), the proportion of individuals who gain more than 7% of weight over the baseline during the first three months is 84% in olanzapine and 64% in risperidone (Correll et al., 2009).

The exact mechanism of antipsychotic-induced weight gain and metabolic disturbance has not been elucidated (Nasrallah, 2008; Newcomer, 2005). Animal studies have indicated that the initial weight gain in individuals using olanzapine is primarily driven by the increases in appetite and food intakes (Davoodi et al., 2009), which is likely caused by the antagonism of histamine H_1_ receptors and 5-HT_2C_ in the presence of dopamine D_2_ receptors (Kirk et al., 2009); whereas others show that risperidone induced weight gain is partly attributable to the alteration of gut microbiome and suppression of energy expenditure (Bahra et al., 2015). In addition, antipsychotics can inhibit the insulin-signaling pathway in the target cells such as muscle cells, hepatocytes and adipocytes from using glucose, and, therefore, cause insulin resistance, which is independent of weight gain and food intake that has been indicated in both patients and healthy controls using antipsychotics (Hahn et al., 2013; van Winkel et al., 2008). Further, the induced obesity can result in high levels of free fatty acids (FFA) and inflammation, which can cause insulin resistance, and antipsychotics can cause direct damage to β-cells, leading to dysfunction and apoptosis of β-cells that increase insulin resistance (Chen et al., 2017).

Antipsychotic-induced metabolic dysfunctions are likely linked to the altered composition of the gut microbiota caused by using antipsychotics. The gut microbiota contains about 500 to 1000 different microorganisms, and the *firmicutes, bacteroidetes, actinobacteria*, and *proteobacteria* are four dominant bacterial phyla in the human gut (Khanna and Tosh, 2014). In germ-free mice, the gut microbiota has been demonstrated as an important environmental factor that regulates energy harvest and storage in the host and plays a critical role in the healthy weight gain and fat composition (Bäckhed et al., 2004; Backhed et al., 2007). Compared with conventionally raised mice, germ-free mice tend to have less total fat and are resistant to diet-induced obesity, which is likely through two complementary, but independent mechanisms of 1) elevated levels of the fasting-induced adipose factor (Fiaf), which induces a key regulator (Pgc-1a) of energy metabolism and 2) increased adenosine monophosphate activated protein kinase (AMPK) activity that increases the metabolism of fatty acids (Backhed et al., 2007). In a double-blind, randomized, placebo-controlled parallel-group clinical trial, glycemic conditions are improved by probiotics and particularly synbiotic supplements in pre-diabetic individuals (Kassaian et al., 2018). In addition, in rats chronically treated with olanzapine, altered fecal microbiota profiles were observed, and they might be associated with metabolic effects induced by olanzapine (Davey et al., 2012). Supplementation of vitamin D plus probiotics also showed clinical and metabolic improvements compared with the placebo in patients with schizophrenia (Ghaderi et al., 2019) who had been treated with chlorpromazine and anticholinergic agents but not concurrently treated with probiotics in the past six months.

The investigations of agents such as antibiotics and probiotics in olanzapine-induced metabolic dysfunctions (Davey et al., 2013), metabolic diseases, and obesity have been encouraging in animal models (Cani et al., 2008; Murphy et al., 2013). Although the number of existing studies about antipsychotics on the gut microbiota is limited, a few studies in rodents have found that olanzapine- and risperidone-induced weight gain and metabolic dysfunction were associated with the changes in the gut microbiota (Bahra et al., 2015; Bretler et al., 2019). However, a minimal number of studies have been conducted in humans (Kang et al., 2019), in particular on antipsychotics-induced metabolic disturbances. A longitudinal study of five male adolescents with psychiatric disorders treated with risperidone showed that a gradual increase in the ratio of *Firmicutes* to *Bacteroidetes* was associated with an increase in body mass index (BMI) compared with antipsychotic-naïve psychiatric controls, and the gut microbiome of the participants was enriched with genes for short-chain fatty acids (SCFA) and serotonin metabolism (Bahr et al., 2015). However, no other outcome measures were evaluated. In this study, we assessed the efficacy of probiotics supplements in preventing olanzapine-induced metabolic disturbance and weight gain in drug-naive first-episode patients with schizophrenia.

## 2. Material and methods

### 2.1 Trial design

This study was a randomized-controlled clinical trial designed to evaluate the efficacy of probiotics for reducing olanzapine-induced metabolic disturbance and weight gain. Participants were randomly assigned (without any restriction or stratification) through a computer-generated random number to the treatment of either olanzapine plus probiotics (olanzapine +) or olanzapine monotherapy in blocks of 4 to ensure approximately equal numbers of participants within the two treatment groups. The treatment assignment (randomization) was determined after the completion of all assessments and acceptance into the study.

The authors assert that all procedures contributing to this work comply with the ethical standards of the relevant national and institutional committees on human experimentation and with the Helsinki Declaration of 1975, as revised in 2008. All procedures involving human patients were approved by the ethics committee of the Second Xiangya Hospital of Central South University (2017-027). Written informed consent was obtained from all patients.

### 2.2 Participants

All participants were recruited (November 30, 2017 to February 01, 2019) by clinical investigators from the Department of Psychiatry of the Second Xiangya Hospital at Central South University, China. Patients were aged 18 to 50 years, the first psychotic episode of schizophrenia and met diagnostic criteria of the Diagnostic and Statistical Manual of Mental Disorders Fifth Edition (DSM-V). All patients were ensured not having used any antipsychotics or recreational drugs at least three months before enrollment and without abnormal in physical examinations, laboratories, and electrocardiogram (ECG) tests.

Exclusions: 1) female patients who were pregnant or lactating at the time of enrollment, 2) patients with mental retardation, addictive disorders, and 3) patients with specific systemic diseases, metabolic disorders, or medical conditions such as diabetes mellitus, dyslipidemia, cardiovascular diseases, and hypertension.

### 2.3 Randomization

Participants were randomly assigned 1:1 to olanzapine+ or olanzapine through a computer-generated random number. A research nurse conducted the randomization. Treating clinicians were not blinded to trial groups, but independent assessors who were blinded to the information on the treatment group evaluated all primary and most of the secondary outcomes.

### 2.4 Intervention

The participants were randomly assigned by investigators to a 12-week treatment of olanzapine+, with olanzapine (15-20mg/day at 8:00 p.m.) plus probiotics (Bifico, triple live bacteria oral capsule, 1680 mg/day, 840mg bid), or olanzapine (15-20mg/day at 8:00 p.m.). Bifico (Shanghai Xinyi Pharmaceutical Inc., Shanghai, China), supplied by the manufacturer, is one kind of commonly used probiotics supplement in China and contains live bacteria of *Bifidobacterium, Lactobacillus* and *Enterococcus* at least 5.0×10^7^ CFU/g. The participants’ adherences to probiotics and olanzapine treatment for each visit was defined as taking more than 80% of the study drug dosage prescribed for that interval. If a participant was non-adherent, both the patient and caregiver were counseled on the importance of taking the prescribed amount of study medication.

### 2.5 Outcome Measures

Primary outcomes were the change in fasting serum insulin, insulin resistance, body mass index (BMI), and weight gain. BMI is calculated as weight in kilograms divided by height in meter squared, and insulin resistance was calculated based on a formula of homeostatic model assessment for insulin resistance (HOMA-IR), by fasting insulin [mIU/L]x fasting glucose [mmol/L]/22.5.

Secondary outcomes were 1) the increase in the levels of lipids, including high-density lipoprotein cholesterol (HDL), low-density lipoprotein cholesterol (LDL), total cholesterol, and triglyceride, 2) Psychopathologic symptoms measured by positive and negative symptom scales (PANSS), and 3) appetite increase. We used a self-reported scale to evaluate appetite. The scale included four items: how hungry you are, how full you are, how strong desire to eat, and how much food you think you can eat. They were measured at a scale of 0-10. A composite score was calculated to measure the overall appetite increase. Compared with baseline, the composite score increased ≥1 was defined as appetite increase, decreased ≥1 was defined as appetite decrease, and an increase or decrease less than 1 was defined as no change in appetite.

### 2.6 Sample size

The sample size was calculated for the primary outcome of HOMA-IR based on the Hotelling-Lawley F test with a linear exponent autoregressive correlation structure (Xu et al., 2020), a base correlation of 0.8, and decay of 0.3 over the time. The total sample size required for a power of 80%, a level of 0.05 with a standard deviation of 0.6 and 0.8 were 44 and 74, respectively; and our actual sample of 76 patients met the calculation.

### 2.7 Data collection

All patients who received the treatment were scheduled to having a clinical evaluation through the scheduled follow-up at week 4, 8, and 12. The baseline assessments included demographics, a comprehensive medical history, physical examination, anthropometric measurements (weight and height), and PANSS score. The laboratory tests at baseline included fasting glucose, lipids and insulin, liver and renal function, blood counts, and electrocardiogram. At each follow-up visit, all baseline clinical evaluations, including physical examination, anthropometric measurements, and laboratory tests, were repeated. The Treatment Emergent Symptom Scale (TESS) (Guy) was used to record adverse events throughout the clinical trial. PANSS were also evaluated at week 12, the end of the trial.

We assessed the appetite half an hour before lunch every day within the first two weeks of the treatments and at each follow-up visit.

### 2.8 Statistical Analysis

Data were examined and processed before the analysis. Summary statistics including mean, median, and standard deviation were calculated for all variables at baseline and each follow-up. Through a comparison of mean and median, one can have a quick assessment of any severe violation of the fair assumption for the statistical methods used for analysis (Zhang and Hughes, 2019). General linear random-effect model(José A. Apud et al., 2012; Wu et al., 2016; Xu et al., 2020) was employed to estimate the fixed-effect of treatments on primary and secondary outcomes over time after adjustment for the baseline characteristics of age, sex, duration of illness, which provide a better analysis in presence of missing or dropouts than the last observation carried forward. The covariance matrix chooses first-order autoregressive [AR (1)] over the time of treatment. Post-hoc least square estimates of mean and standard errors were obtained for graphic presentation; correction for multiple testing was performed using Dunnett’s test to compare the outcomes at each follow-up visit with the baselines. The analysis was performed using SPSS and SAS/STAT 13.2; a two-sided level of 0.05 was considered as statistically significant.

Ancillary analyses were also performed on the four primary outcomes while adjusting for the status of appetite increase, which was coded as a binary outcome. The main effect of appetite increase, plus the interaction between appetite increase and treatment time was included in the regression model to assess the treatment effect over time. The report of the study followed the CONSORT 2010 Statement updated guidelines for reporting parallel group randomized trials and related recommendations for reporting of clinical trials as well as calls for transparency in the report for clinical and preclinical research (Landis et al., 2012; Zhang and Hughes, 2019).

## 3. Results

Participants were recruited from November 30, 2017, to February 01, 2019. Of 126 patients screened for eligibility (**Figure 1**), 17 patients did not meet the inclusion or met exclusion criteria, 19 patients declined to participate; 90 patients were randomly and equally assigned to the group of olanzapine+ or olanzapine monotherapy. During the trial period, 14 patients did not receive intervention with medication as randomized or withdrew the consent; so, 76 patients were evaluated with follow-up to collect the outcome measurements, which included 39 patients in the olanzapine+ and 37 in the olanzapine, who were followed at least one time. Finally, 88.2% of patients completed the 12-week treatment. We examined the nine individuals who did not complete the follow-up at week 8 or 12, found no significant difference in baseline characteristics of patients from those patients who completed all follow-ups (**Table S1**).

**Figure 1.**
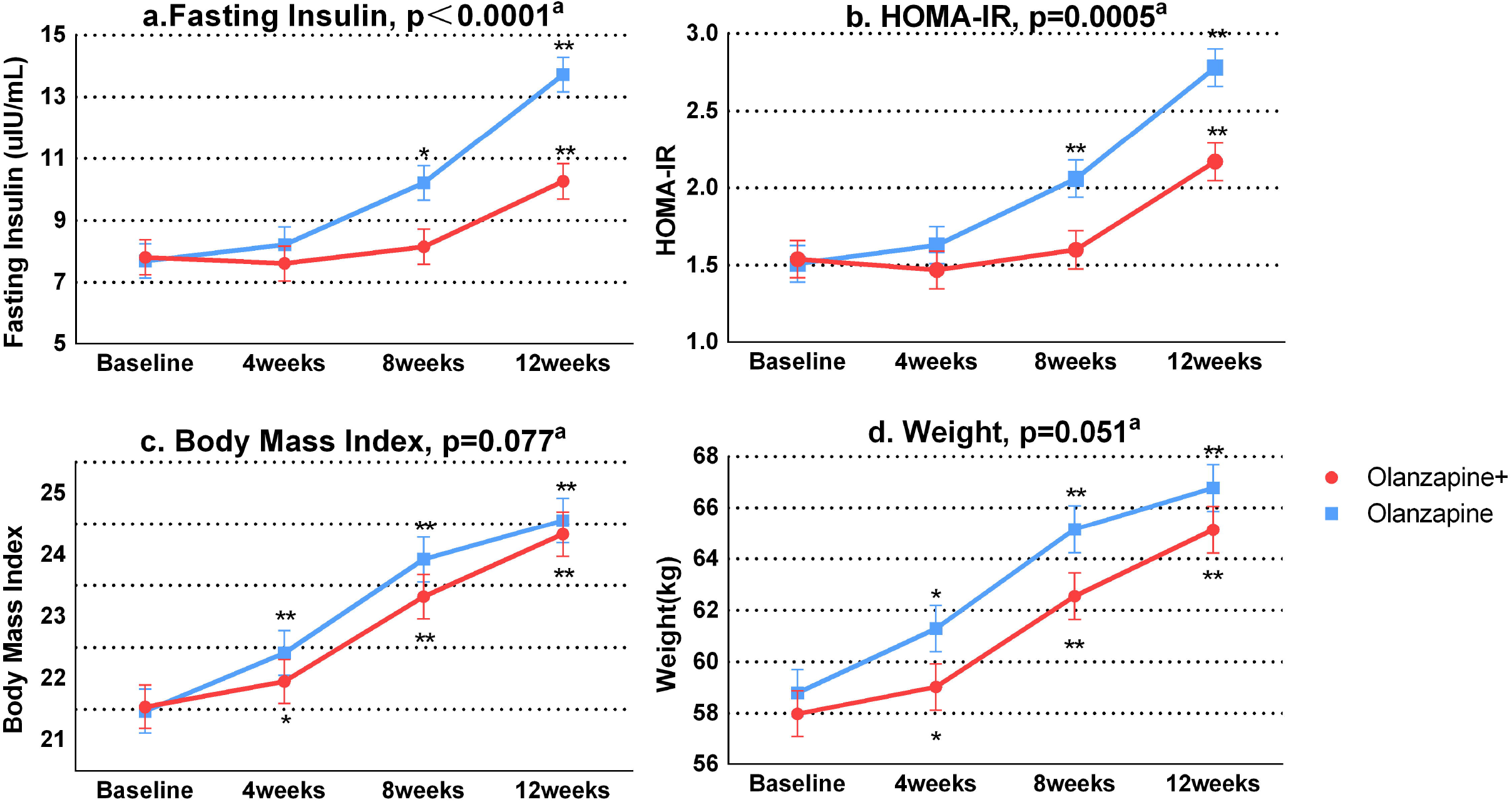
Flowchart of Patients in Randomized Trial of Probiotic Supplements for Olanzapine-induced Metabolic Disturbances and Weight Gain

**Table 1** presents the demographics and characteristics of the patients by treatment group. There was no significant difference in all the variables between groups, which were expected due to the randomized assignments. About 65-72% of the patients were males, and 35-36% were cigarette smokers. The mean age was 24 years, and the duration of illness was about 11 months; the total PANSS score was about 80. The baseline characteristics and outcome measures were not different (*p*>0.05) between two treatments except for a slight difference in LDL (*p*=0.0017) and total cholesterol (*p*=0.037), which were higher in the olanzapine+ than in the olanzapine monotherapy. The higher level of LDL and total cholesterol at baseline assured at least no over-estimation of the interventional effect of probiotic supplements on the secondary outcomes when olanzapine+ are assumed to reduces the metabolic disturbance of lipid profiles. The summary statistics of the key outcomes at baseline and follow-up time points by treatment group were presented in the supplementary (**Table S2**)

**Table 1.**
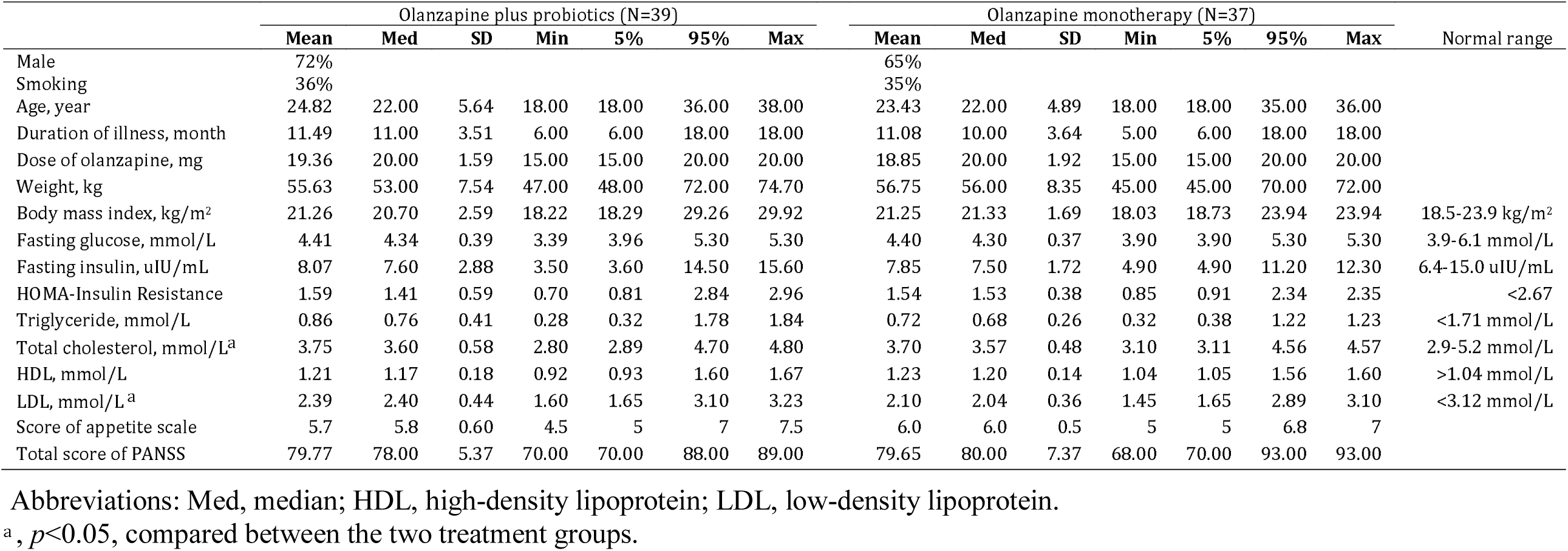
Baseline Characteristics of the Patients by Treatment Group

|  | Olanzapine plus probiotics (N=39) |  |  |  |  |  |  | Olanzapine monotherapy (N=37) |  |  |  |  |  |  | Normal range |
| --- | --- | --- | --- | --- | --- | --- | --- | --- | --- | --- | --- | --- | --- | --- | --- |
|  | Mean | Med | SD | Min | 5% | 95% | Max | Mean | Med | SD | Min | 5% | 95% | Max |  |
| Male | 72% |  |  |  |  |  |  | 65% |  |  |  |  |  |  |  |
| Smoking | 36% |  |  |  |  |  |  | 35% |  |  |  |  |  |  |  |
| Age, year | 24.82 | 22.00 | 5.64 | 18.00 | 18.00 | 36.00 | 38.00 | 23.43 | 22.00 | 4.89 | 18.00 | 18.00 | 35.00 | 36.00 |  |
| Duration of illness, month | 11.49 | 11.00 | 3.51 | 6.00 | 6.00 | 18.00 | 18.00 | 11.08 | 10.00 | 3.64 | 5.00 | 6.00 | 18.00 | 18.00 |  |
| Dose of olanzapine, mg | 19.36 | 20.00 | 1.59 | 15.00 | 15.00 | 20.00 | 20.00 | 18.85 | 20.00 | 1.92 | 15.00 | 15.00 | 20.00 | 20.00 |  |
| Weight, kg | 55.63 | 53.00 | 7.54 | 47.00 | 48.00 | 72.00 | 74.70 | 56.75 | 56.00 | 8.35 | 45.00 | 45.00 | 70.00 | 72.00 |  |
| Body mass index, kg/m<sup>2</sup> | 21.26 | 20.70 | 2.59 | 18.22 | 18.29 | 29.26 | 29.92 | 21.25 | 21.33 | 1.69 | 18.03 | 18.73 | 23.94 | 23.94 | 18.5-23.9 kg/m<sup>2</sup> |
| Fasting glucose, mmol/L | 4.41 | 4.34 | 0.39 | 3.39 | 3.96 | 5.30 | 5.30 | 4.40 | 4.30 | 0.37 | 3.90 | 3.90 | 5.30 | 5.30 | 3.9-6.1 mmol/L |
| Fasting insulin, uIU/mL | 8.07 | 7.60 | 2.88 | 3.50 | 3.60 | 14.50 | 15.60 | 7.85 | 7.50 | 1.72 | 4.90 | 4.90 | 11.20 | 12.30 | 6.4-15.0 uIU/mL |
| HOMA-Insulin Resistance | 1.59 | 1.41 | 0.59 | 0.70 | 0.81 | 2.84 | 2.96 | 1.54 | 1.53 | 0.38 | 0.85 | 0.91 | 2.34 | 2.35 | <2.67 |
| Triglyceride, mmol/L | 0.86 | 0.76 | 0.41 | 0.28 | 0.32 | 1.78 | 1.84 | 0.72 | 0.68 | 0.26 | 0.32 | 0.38 | 1.22 | 1.23 | <1.71 mmol/L |
| Total cholesterol, mmol/L<sup>a</sup> | 3.75 | 3.60 | 0.58 | 2.80 | 2.89 | 4.70 | 4.80 | 3.70 | 3.57 | 0.48 | 3.10 | 3.11 | 4.56 | 4.57 | 2.9-5.2 mmol/L |
| HDL, mmol/L | 1.21 | 1.17 | 0.18 | 0.92 | 0.93 | 1.60 | 1.67 | 1.23 | 1.20 | 0.14 | 1.04 | 1.05 | 1.56 | 1.60 | >1.04 mmol/L |
| LDL, mmol/L<sup>a</sup> | 2.39 | 2.40 | 0.44 | 1.60 | 1.65 | 3.10 | 3.23 | 2.10 | 2.04 | 0.36 | 1.45 | 1.65 | 2.89 | 3.10 | <3.12 mmol/L |
| Score of appetite scale | 5.7 | 5.8 | 0.60 | 4.5 | 5 | 7 | 7.5 | 6.0 | 6.0 | 0.5 | 5 | 5 | 6.8 | 7 |  |
| Total score of PANSS | 79.77 | 78.00 | 5.37 | 70.00 | 70.00 | 88.00 | 89.00 | 79.65 | 80.00 | 7.37 | 68.00 | 70.00 | 93.00 | 93.00 |  |

Abbreviations: Med, median; HDL, high-density lipoprotein; LDL, low-density lipoprotein.
<sup>a</sup>, $p<0.05$ , compared between the two treatment groups.

### 3.1 Primary outcomes

Figure 2. provides an intuitive display of the treatment effects over time. The addition of probiotic supplements concurrently with olanzapine medication significantly slowed the increase in fasting insulin (*p*<0.0001) and HOMA-IR (*p*=0.0005), but only had a slight effect on reducing the increase in BMI (*p*=0.077) and weight gain (*p*=0.051).

**Table 2** provides the detailed estimates of treatment effect. Specifically, patients received treatment of olanzapine+ had an increase in the levels of fasting insulin by 2.45 uIU/mL, from 7.81 at baselines to 10.26 at week 12 (*p*<0.0001), which was much lower than an increase by 6.03 uIU/mL (p<0.0001), from 7.69 at baselines to 13.72 at week 12 in the olanzapine monotherapy, and there was a significant difference in the level of fasting insulin at the end of the trial between two groups (difference, −3.46, 95% CI [-5.0,-1.91]; *p*<0.0001). Similarly, the HOMA-IR increased by 0.63 (*p<*0.0001), from 1.54 at the baseline to 2.17 at week 12 in the olanzapine+ group, which was lower than the increase by 1.27 (*p*<0.0001), from 1.51 at the baselines to 2.78 at week 12 in the olanzapine monotherapy; also, there was a significant difference in the HOMA-IR at the end of the trial between two groups (difference, −0.61, 95% CI [-0.94,-0.28]; *p*=0.0004). Both BMI and body weight significantly increased at week 4 compared with baseline and continued to increase throughout the study in both the olanzapine+ and olanzapine group, but overall, these increases were not significantly different between the two treatment groups (*p*>0.05).

**Table 2.** Estimates of Treatment Effect on Primary Outcomes by Group

|  | Olanzapine+ |  |  |  | Olanzapine |  |  |  | Group difference |  |  |  |  |
| --- | --- | --- | --- | --- | --- | --- | --- | --- | --- | --- | --- | --- | --- |
|  | N | LS Mean | SE | *P<sub>adj</sub>* | N | LS mean | SE | *P<sub>adj</sub>* | Beta | SE | 95%CI | *P* | *P<sup>a</sup>* |
| Fasting Insulin, uIU/mL |  |  |  |  |  |  |  |  |  |  |  |  |  |
| Baseline | 35 | 7.81 | 0.564 | ref | 36 | 7.69 | 0.553 | ref | 0.119 | 0.772 | -1.394, 1.632 | 0.8775 | <.0001 |
| 4 Weeks | 35 | 7.61 | 0.564 | 0.7352 | 34 | 8.22 | 0.562 | 0.6315 | -0.612 | 0.78 | -2.141, 0.917 | 0.434 |  |
| 8 Weeks | 33 | 8.15 | 0.575 | 0.5475 | 34 | 10.21 | 0.562 | 0.0012 | -2.061 | 0.788 | -3.605, -0.517 | 0.0098 |  |
| 12 Weeks | 33 | 10.26 | 0.575 | <.0001 | 34 | 13.72 | 0.562 | <.0001 | -3.455 | 0.788 | -4.999, -1.911 | <.0001 |  |
| HOMA-IR |  |  |  |  |  |  |  |  |  |  |  |  |  |
| Baseline | 35 | 1.54 | 0.121 | ref | 36 | 1.51 | 0.118 | ref | 0.037 | 0.165 | -0.286, 0.360 | 0.8254 | 0.0005 |
| 4 Weeks | 35 | 1.47 | 0.121 | 0.5875 | 34 | 1.63 | 0.12 | 0.577 | -0.151 | 0.167 | -0.478, 0.176 | 0.3665 |  |
| 8 Weeks | 33 | 1.6 | 0.123 | 0.8024 | 34 | 2.06 | 0.12 | 0.0007 | -0.459 | 0.169 | -0.790, -0.128 | 0.0073 |  |
| 12 Weeks | 33 | 2.17 | 0.123 | <.0001 | 34 | 2.78 | 0.12 | <.0001 | -0.611 | 0.169 | -0.942, -0.280 | 0.0004 |  |
| Body mass index, kg/m<sup>2</sup> |  |  |  |  |  |  |  |  |  |  |  |  |  |
| Baseline | 39 | 21.54 | 0.355 | ref | 37 | 21.47 | 0.359 | ref | 0.068 | 0.493 | -0.898, 1.034 | 0.8905 | 0.077 |
| 4 Weeks | 39 | 21.95 | 0.355 | 0.0036 | 37 | 22.41 | 0.359 | <.0001 | -0.465 | 0.493 | -1.431, 0.501 | 0.3488 |  |
| 8 Weeks | 37 | 23.32 | 0.357 | <.0001 | 34 | 23.92 | 0.362 | <.0001 | -0.6 | 0.497 | -1.574, 0.374 | 0.2308 |  |
| 12 Weeks | 33 | 24.33 | 0.361 | <.0001 | 34 | 24.55 | 0.362 | <.0001 | -0.217 | 0.5 | -1.197, 0.763 | 0.6649 |  |
| Weight, kg |  |  |  |  |  |  |  |  |  |  |  |  |  |
| Baseline | 39 | 57.98 | 0.897 |  | 37 | 58.79 | 0.907 |  | -0.817 | 1.247 | -3.261, 1.627 | 0.514 | 0.051 |
| 4 Weeks | 39 | 59.02 | 0.897 | 0.0035 | 37 | 61.29 | 0.907 | 0.0035 | -2.273 | 1.247 | -4.717, 0.171 | 0.0718 |  |
| 8 Weeks | 37 | 62.55 | 0.903 | <.0001 | 34 | 65.16 | 0.915 | <.0001 | -2.608 | 1.257 | -5.072, -0.144 | 0.0409 |  |
| 12 Weeks | 33 | 65.14 | 0.913 | <.0001 | 34 | 66.78 | 0.915 | <.0001 | -1.64 | 1.264 | -4.117, 0.837 | 0.1976 |  |

Abbreviations: Olanzapine+ , olanzapine plus probiotics; HOMA-IR, HOMA insulin resistance.
LS mean, post-hoc least square estimate of mean.
*P<sup>a</sup>* , *p* value for testing the difference in insulin and body mass index over time between treatments.
*P<sub>adj</sub>*, *p* value for testing the metabolic change versus the baseline within the treatment group.

**Figure 2.**
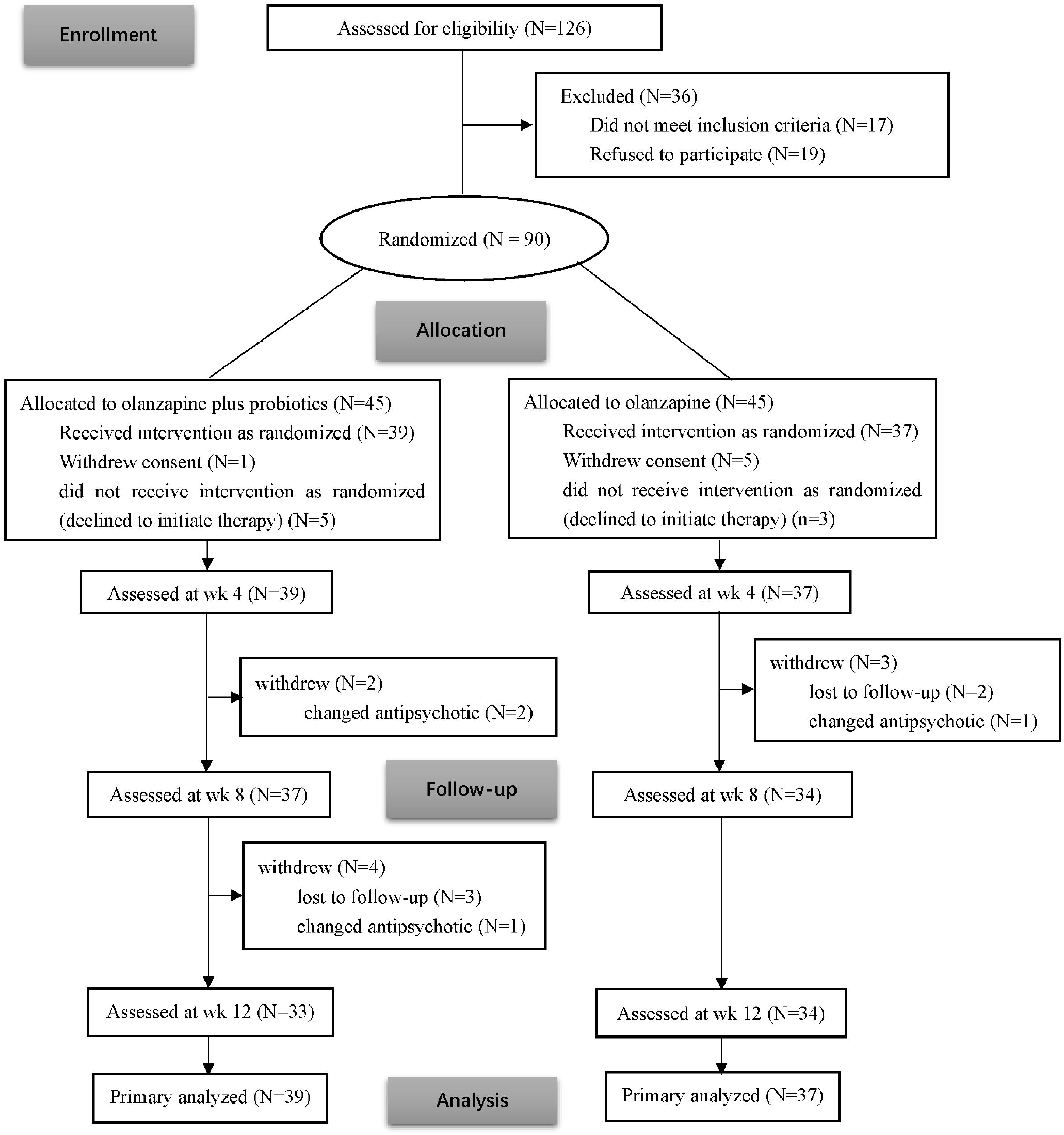
The Estimated Effect of Probiotic supplements on Olanzapine-induced Elevation of Fasting Insulin, Insulin Resistance, Body Mass Index, and Weight Plots were presented with LS mean, and SEM; Body mass index, which is calculated as weight in kilograms divided by height in meters squared; HOMA-IR, Homeostatic Model Assessment of Insulin Resistance, which is calculated as insulin level (mIU/L)×fasting glucose(mmol/L)/22.5; ^a^ P value for testing the difference in outcomes over time between treatments.

### 3.2 Secondary outcomes

#### Appetite increase

Olanzapine plus probiotic supplements slightly reduced appetite increase. In the olanzapine+ group, 38.5% (15/39) of patients had appetite increase, which was lower than 56.8% (21/37) in the olanzapine group (log-rank test, *p*=0.0666; likelihood ratio test, *p*=0.0255) (**Figure S1)**. The difference in the appetite increase became no significant (*p*=0.1211) between two treatments after adjustment for sex, age, duration of illness, and dosage of olanzapine taken in the Cox regression analysis. However, on average, patients treated with olanzapine+ were about 40% less likely (Hazard Ratio, HR=0.589) to have an increase in appetite than those who received olanzapine monotherapy (**Table S3**).

#### Psychopathological symptoms

The psychopathological symptoms improved in both groups after treatment for 12 weeks but the improvement was not significantly different between groups. The PANSS total score was reduced by 24.5 (*p*<0.0001) from 80.18 at the baseline to 55.67, after olanzapine+ for 12 weeks and by 22.7 (*p*<0.0001), from 79.5 at the baseline to 56.8, after olanzapine treatment. However, the improvement of PANSS total score was not significantly different between two groups (*p*=0.48). We also observed a similar pattern in positive, negative, and the general psychopathology (**Table S4**).

#### Lipid profiles

The lipid profiles that comprised triglyceride, total cholesterol, HDL, and LDL increased very significantly (*p*<0.0001) in both the olanzapine+ and the olanzapine group, but no significant difference in triglyceride (*p*=0.21), HDL (*p*=0.60) and LDL (*p*=0.55), except total cholesterol (*p*=0.028) between two treatment groups (**Table S5**).

### 3.3 Appetite modulated antipsychotics-induced metabolic change

While the addition of probiotic supplements slightly reduced the likelihood and timing of the change in appetite, which had a significant effect on the metabolic disturbance in both groups. Patients with appetite increase had a significant (*p*<0.0001) elevation of all primary outcomes of fasting insulin, HOMA-IR, BMI, and weight than those patients without appetite increase (**Table 3**). The metabolic disturbance increased gradually with the duration of treatment and became significant at week 12 in BMI (*p*=0.028), fasting insulin (*p*<0.0001), and HOMA-IR (*p*<0.0001). However, we noted that the significant differences between two groups in weight gain were primarily driven by the baseline measurements, in which the weight was significantly different (*p*=0.0065) at baseline but no difference after treatment at week 12 (*p*=0.28). Of note, appetite also modulated the antipsychotic-induced changes in the secondary outcomes such as triglyceride (*p*=0.019), total cholesterol (*p*=0.0002), and LDL (*p*<0.0001), but not in HDL (*p*=0.054).

**Table 3.**
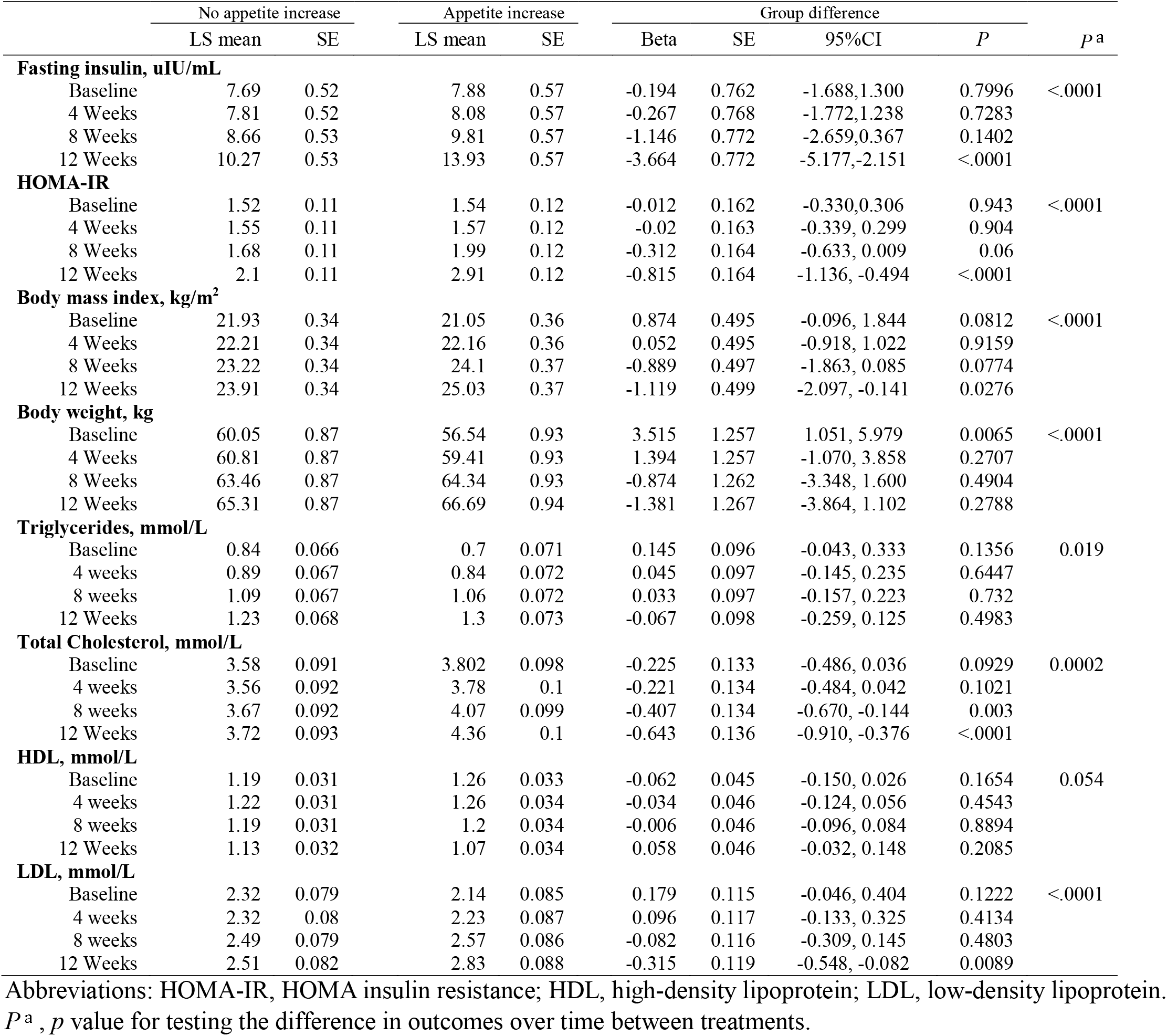
Appetite Modulated Metabolic Disturbance

### 3.4 Ancillary analysis

We further investigated if the effect of probiotic supplements on the metabolic disturbance was through the less increase in appetite (**Table S6**). After adjustment for the status of appetite increase, probiotic supplements still significantly reduced the increase in fasting insulin (*p*<0.0001) and HOMA-IR (*p*=0.0012); the effect also started at week 8 after treatment, suggesting that the impact of probiotic supplements on the increase in fasting insulin and HOMA-IR were mostly independent of appetite increase. However, two treatments had no significant impact on the change in BMI (*p*=0.143) and body weight (*p*=0.11) during the trial after adjustment for appetite increase.

### 3.5 Adverse events

Adverse events considered grade 3 or higher that were reported at least 5% of the evaluable patients (**Table 4**). The most common adverse events in both groups were hypoactivity (26-30%), somnolence (26-27%), and abnormal in liver function test (15-16%), but they were not significantly different between the two treatment groups. The only adverse event reported as possibly related to the probiotic supplements was diarrhea, which was 10% in the olanzapine+ but only 3% in the olanzapine group. In contrast, constipation was 3% in olanzapine+, but 14% in the olanzapine monotherapy. None of these was suitable for pursuing a formal statistical test due to small sample size.

**Tables 4.** Common Adverse Events (>5%) in the two Treatment Groups a

*P*<sup>a</sup>, *p* value for testing the difference in outcomes over time between treatments. **Tables 4** Common Adverse Events (>5%) in the two Treatment Groups <sup>a</sup>
|  | Olanzapine plus probiotic<br>(N=39) |  | Olanzapine monotherapy<br>(N=37) |  |
| --- | --- | --- | --- | --- |
| **Adverse events** | N | % | N | % |
| Hypoactivity | 10 | 26 | 11 | 30 |
| Somnolence | 10 | 26 | 10 | 27 |
| Abnormal in liver function test | 6 | 15 | 6 | 16 |
| Akathisia | 2 | 5 | 3 | 8 |
| Constipation | 1 | 3 | 5 | 14 |
| Diarrhea | 4 | 10 | 1 | 3 |
| Dizzy | 2 | 5 | 3 | 8 |
| Weight gain (7% from baseline) | 24 | 62 | 27 | 73 |

<sup>a</sup>, Assessment was based on 76 patients who took at least one dose of olanzapine or olanzapine plus probiotics and were evaluable for adverse event assessment.

## 4. Discussion

The main findings of this trial were that probiotic supplements with olanzapine attenuated antipsychotics-induced elevation of fasting serum insulin and insulin resistance as measured by HOMA-IR while retaining the treatment effect on psychopathological symptoms. However, we did not find a significant benefit on BMI and weight gain, nor the secondary outcomes of appetite increase as well as lipid profiles. Our study provided the first evidence for probiotic supplements in preventing antipsychotics-induced insulin resistance in patients with drug-naive first-episode schizophrenia while concurrently using antipsychotics.

Our findings that probiotic supplements reduced the olanzapine-induced increase in fasting insulin and insulin resistance were consistent with previous multiple studies in individuals with diabetic conditions. 1) One randomized, double-blind, placebo-controlled trial in Thailand finds that supplementation of probiotics containing *Bifidobacterium* (*B*.) and *Lactobacillus* (*L*.) daily for four consecutive weeks improves the plasma fasting insulin and HOMA-IR in women diagnosed with gestational diabetes mellitus (GDM) between 24–28 weeks (Kijmanawat et al., 2019), compared to the placebo group; 2) In another randomized clinical trial (RCT) in Ukraine, supplementation of multi-probiotics, including concentrated biomass of 14 probiotic bacteria genera *Bifidobacterium, Lactobacillus, Lactococcus, Propionibacterium* for eight weeks, was associated with the reduction of HOMA-IR in individuals with type 2 diabetes (Kobyliak et al., 2018). Both trials used probiotics that contain *Bifidobacterium* and *Lactobacillus*, two common bacteria for probiotics, as that used in our study; 3) A meta-analysis of six RCTs involved 830 patients also confirmed that supplementation of probiotics significantly reduced the serum fasting insulin and HOMA-IR in women with GDM (Pan et al., 2019). In addition, in a double blind, randomized, placebo-controlled parallel-group clinical trial for 24 weeks, glycemic conditions can be improved significantly by probiotics, particularly synbiotic (prebiotic plus probiotic) supplements in pre-diabetic individuals (Kassaian et al., 2018). Insulin resistance plays a crucial role in the olanzapine-induced metabolic disturbance, including lipid profiles and weight gain (Wu et al., 2016; Wu et al., 2008), which at least partly depends on appetite increase (Davoodi et al., 2009). However, we found that the benefit of probiotic supplements on preventing the olanzapine-induced insulin resistance was still significant regardless of appetite increase.

Our study did not find a significant effect of probiotic supplements on the lipid profiles, although reports have shown that probiotics have a significant effect on lipid metabolism. In the most recent meta-analysis of ten RCTs with 1,139 participants, probiotics supplements were shown to have improved effect on glucose, HOMA-IR, and lipid metabolism in pregnant women (Han et al., 2019). A meta-analysis of 13 RCTs with 485 patients with high, borderline high, or healthy cholesterol also shows that a diet rich in probiotics decreases the total cholesterol and LDL cholesterol concentration in plasma (Guo et al., 2011). In addition, an RCT study of vitamin D plus probiotic supplements (8×10^9^ CFU), containing *B*.*bifidum, L*.*acidophilus, L*.*reuteri*, and *L*.*fermentum* (each 2 × 10^9^) for 12 weeks in patients with chronic schizophrenia who had used but not concurrently with antipsychotics, showed a significant effect on the improvement of metabolic profiles including fasting glucose, fasting insulin and HOMA-IR, triglycerides and total cholesterol levels (Ghaderi et al., 2019). The limited effect of probiotic supplements on lipid profiles could be due to the short duration of treatment in this trial because our previous trials of metformin on antipsychotic-induced dyslipidemia in schizophrenia indicate that the treatment effect on lipids requires a prolonged duration of treatment (24 weeks) than on insulin or insulin resistance (12 weeks) (Wu et al., 2016).

Increasing evidence shows that there is a strong metabolic risk intrinsic to schizophrenia even in the absence of antipsychotic drug treatment (Freyberg et al., 2017). As in this study with drug-naive patients, we noted elevated levels of lipids in one of the groups. This would make a study of definitive mechanisms more challenging for antipsychotic-induced metabolic disturbance. There have been studies indicating that genetic variants at *TCF7L2* and *PSMD9* were independently associated with schizophrenia and type 2 diabetes (Freyberg et al., 2017). In addition, the central nervous system (e.g. hypothalamus) where the neurotransmitter and neuropeptide system such as dopamine, serotonin, and histamine may play a role in the intrinsic metabolic risk. This is because that almost all antipsychotics act on dopamine receptors (D2 and D3) and/or serotonin receptors, and polymorphism in *DRD2, DRD3, 5HT2a*, and *5HT2c* have been associated with both antipsychotic-induced metabolic dysfunction and schizophrenia (Freyberg et al., 2017). In fact, some polymorphisms in *TCF7L2* and *DRD2* have been found to be shared by schizophrenia, bipolar, and major depressive disorders (Xia et al., 2019). The effect of probiotic supplements on the levels of fast insulin and insulin resistance may support that the gut-brain axis plays a role in modulating antipsychotic-induced metabolic dysfunction and might have implications for reducing the risk of schizophrenia at least for a partial population of the patients with schizophrenia.

To our knowledge, this was the first study to assess probiotic supplements on preventing olanzapine-induced metabolic disturbance and weight gain in drug naïve first-episode patients with schizophrenia. Supplementation of probiotics was in favor of preventing olanzapine-induced elevation of fasting serum insulin and insulin resistance, and was shown to be well tolerated and safe in the participants.

### 4.1 Limitation

This study had some limitations. First, we did not use a placebo in this trial, although the treatment information was blinded to the clinical evaluators. Second, this was a 12-week trial, and about 20% of patients were lost to follow-up due to the inability to take the medication as a randomized assignment. In such a limited duration of treatment, we did not know whether the improved insulin sensitivity could sustain after patients stopped taking probiotic supplements. Third, this was a single-center clinical trial, which had the advantage of homogenous samples but may not allow us to address the variability across regions. Therefore, it may limit the generalizability of the study results to the study population in a south-central province of China. In addition, some related measurements such as adiposity and leptin levels were not measured; we did not examine the microbiome that would strengthen the study and help reveal the underlying mechanism of treatment response. These may require additional effort such as funding support and development of study protocols.

### 4.2 Generalizability

Despite some limitations, our study provided the first evidence that probiotic supplements can be used for preventing antipsychotic-induced metabolic disturbance in particular elevation of fasting insulin and insulin resistance. Given that probiotic supplements were generally safe for use, as shown in this trial, the study results may have implications for clinical practice, the supplementations of probiotics can be recommended for concurrent use with antipsychotic treatment in patients with schizophrenia in the study population in the area where the study was conducted. In addition, further research is needed to assess the effect of the probiotic supplements on antipsychotic-induced weight gain and metabolic disturbance in a longer-term and potentially synergistic effect on antipsychotic response.

## Data Availability

The data that support the findings of this study are available from the corresponding author, Renrong Wu, upon reasonable request.

## Acknowledgement

We would like to thank all the participants and researchers who have been involved in the research.

