## Supplemental table for "Probiotic supplements reduce antipsychotic-induced metabolic disturbances in drug-naïve first-episode schizophrenia"

**Affiliation/address:**

1. Psychiatry Department and Mental Health Institute of the Second Xiangya Hospital, Central South University; China National Clinical Research Center on Mental Disorders; China National Technology Institute on Mental Disorders; Hunan medical center for mental health; Hunan Key Laboratory of Psychiatry and Mental Health, Changsha, Hunan 410011, China
2. Beijing Huilongguan Hospital and Peking University Huilongguan Clinical Medical School, Beijing, China
3. Global Clinical and Translational Research Institute, Bethesda, MD, USA
4. Psychiatric Institute, University of Illinois, Chicago, IL, USA
5. Shanghai Institutes for Biological Sciences, Chinese Academy of Sciences, Shanghai 200031, China

\# Authors contributed equally to this work.

**Table S1.** Baseline comparisons between completers and non-completers due to lost to follow-up^a^

|  | Completers (N=67) | | | | |  | Non-completers (N=9) | | | | |
| --- | --- | --- | --- | --- | --- | --- | --- | --- | --- | --- | --- |
|  | Mean | M | SD | Min | Max |  | Mean | M | SD | Min | Max |
| Male | 68.7% |  |  |  |  |  |  | 66.7% |  |  |  |
| Smoking | 35.8% |  |  |  |  |  |  | 44.4% |  |  |  |
| Age, year | 23.9 | 22 | 5.2 | 18.0 | 36.0 |  | 26.1 | 26.0 | 5.9 | 18.0 | 38.0 |
| Duration of illness, month | 11.6 | 12 | 3.6 | 5.0 | 18.0 |  | 9.0 | 9.0 | 1.5 | 7.0 | 12.0 |
| Dose of olanzapine, mg | 19.0 | 20.0 | 1.8 | 15.0 | 20.0 |  | 19.7 | 20 | 0.8 | 17.5 | 20.0 |
| Weight, kg | 55.6 | 53.2 | 7.6 | 45.0 | 72.0 |  | 60.5 | 57.7 | 9.5 | 47.0 | 74.7 |
| Body mass index, kg/m^2^ | 21.1 | 21 | 1.7 | 18.0 | 25.5 |  | 22.6 | 20.7 | 4.3 | 18.8 | 29.9 |
| Fasting glucose, mmol/L | 4.4 | 4.3 | 0.4 | 3.4 | 5.3 |  | 4.3 | 4.2 | 0.3 | 3.9 | 4.7 |
| Fasting insulin, uIU/mL | 8.1 | 7.8 | 2.3 | 3.5 | 15.6 |  | 5.9 | 5.8 | 1.6 | 4.1 | 7.6 |
| HOMA-Insulin Resistance | 1.6 | 1.5 | 0.5 | 0.7 | 3.0 |  | 1.1 | 1.1 | 0.3 | 0.8 | 1.5 |
| Triglyceride, mmol/L | 0.8 | 0.7 | 0.3 | 0.3 | 1.5 |  | 1.0 | 0.8 | 0.5 | 0.5 | 1.8 |
| Total cholesterol, mmol/L | 3.7 | 3.6 | 0.5 | 2.8 | 4.7 |  | 3.8 | 3.6 | 0.6 | 2.9 | 4.8 |
| HDL, mmol/L | 1.2 | 1.2 | 0.2 | 1.0 | 1.7 |  | 1.1 | 1.1 | 0.1 | 0.9 | 1.3 |
| LDL, mmol/L | 2.3 | 2.2 | 0.4 | 1.5 | 3.1 |  | 2.2 | 2.0 | 0.5 | 1.7 | 3.2 |
| Total score of PANSS | 79.3 | 78 | 6.4 | 68.0 | 93.0 |  | 83.0 | 82 | 5.4 | 77.0 | 93.0 |

^a^ There is no significant difference in all baseline characteristics between;

Non-completer, defined as individuals who did not complete all follow-up evaluation.

**Table S2**. Descriptive statistics of primary and secondary metabolic outcomes at baseline and follow-up time point by treatment groups

|  |  | |  | Olanzapine plus probiotics | | |  |  | Olanzapine only | | |
| --- | --- | --- | --- | --- | --- | --- | --- | --- | --- | --- | --- |
|  | Week | | N | Mean | Media | SD |  | N | Mean | Media | SD |
| Weight, kg | | |  |  |  |  |  |  |  |  |  |
|  | | 0 | 39 | 55.63 | 53.00 | 7.54 |  | 37 | 56.75 | 56.00 | 8.35 |
|  | | 4 | 39 | 56.67 | 54.00 | 7.32 |  | 37 | 59.25 | 56.90 | 8.34 |
|  | | 8 | 37 | 59.93 | 57.40 | 6.83 |  | 34 | 63.29 | 62.60 | 7.85 |
|  | | 12 | 33 | 61.54 | 59.80 | 6.10 |  | 34 | 64.91 | 65.75 | 7.87 |
| Body mass index, kg/m^2^ | | | |  |  |  |  |  |  |  |  |
|  | | 0 | 39 | 21.26 | 20.70 | 2.59 |  | 37 | 21.25 | 21.33 | 1.69 |
|  | | 4 | 39 | 21.67 | 21.33 | 2.56 |  | 37 | 22.19 | 22.14 | 1.57 |
|  | | 8 | 37 | 22.99 | 22.95 | 2.56 |  | 34 | 23.85 | 24.11 | 1.64 |
|  | | 12 | 33 | 23.60 | 23.42 | 2.02 |  | 34 | 24.47 | 25.02 | 1.79 |
| Fasting glucose, mmol/L | | | |  |  |  |  |  |  |  |  |
|  | | 0 | 39 | 4.41 | 4.34 | 0.39 |  | 37 | 4.40 | 4.30 | 0.37 |
|  | | 4 | 37 | 4.34 | 4.20 | 0.45 |  | 35 | 4.48 | 4.48 | 0.38 |
|  | | 8 | 37 | 4.43 | 4.30 | 0.38 |  | 34 | 4.53 | 4.55 | 0.36 |
|  | | 12 | 33 | 4.70 | 4.70 | 0.44 |  | 34 | 4.58 | 4.55 | 0.33 |
| Fasting insulin, uIU/mL | | | |  |  |  |  |  |  |  |  |
|  | | 0 | 35 | 8.07 | 7.60 | 2.88 |  | 36 | 7.85 | 7.50 | 1.72 |
|  | | 4 | 35 | 7.86 | 7.20 | 2.65 |  | 34 | 8.42 | 8.40 | 2.01 |
|  | | 8 | 33 | 8.50 | 7.90 | 2.81 |  | 34 | 10.42 | 10.75 | 3.22 |
|  | | 12 | 33 | 10.61 | 10.50 | 3.60 |  | 34 | 13.92 | 13.85 | 5.46 |
| HOMA-Insulin Resistance | | | |  |  |  |  |  |  |  |  |
|  | | 0 | 35 | 1.59 | 1.41 | 0.59 |  | 36 | 1.54 | 1.53 | 0.38 |
|  | | 4 | 35 | 1.52 | 1.38 | 0.57 |  | 34 | 1.67 | 1.62 | 0.42 |
|  | | 8 | 33 | 1.66 | 1.55 | 0.56 |  | 34 | 2.11 | 2.02 | 0.70 |
|  | | 12 | 33 | 2.24 | 2.10 | 0.89 |  | 34 | 2.83 | 2.82 | 1.12 |
| Triglyceride, mmol/L | | | |  |  |  |  |  |  |  |  |
|  | | 0 | 39 | 0.86 | 0.76 | 0.41 |  | 37 | 0.72 | 0.68 | 0.26 |
|  | | 4 | 37 | 0.89 | 0.78 | 0.37 |  | 35 | 0.87 | 0.87 | 0.34 |
|  | | 8 | 37 | 1.11 | 1.09 | 0.40 |  | 34 | 1.08 | 1.00 | 0.44 |
|  | | 12 | 33 | 1.25 | 1.25 | 0.36 |  | 34 | 1.26 | 1.18 | 0.54 |
| Total cholesterol, mmol/L | | | |  |  |  |  |  |  |  |  |
|  | | 0 | 39 | 3.75 | 3.60 | 0.58 |  | 37 | 3.70 | 3.57 | 0.48 |
|  | | 4 | 37 | 3.84 | 3.76 | 0.54 |  | 35 | 3.56 | 3.42 | 0.49 |
|  | | 8 | 37 | 3.97 | 4.02 | 0.63 |  | 34 | 3.84 | 3.88 | 0.55 |
|  | | 12 | 33 | 4.02 | 4.07 | 0.60 |  | 34 | 4.08 | 4.15 | 0.79 |
| HDL, mmol/L | | |  |  |  |  |  |  |  |  |  |
|  | | 0 | 39 | 1.21 | 1.17 | 0.18 |  | 37 | 1.23 | 1.20 | 0.14 |
|  | | 4 | 37 | 1.25 | 1.21 | 0.21 |  | 35 | 1.23 | 1.21 | 0.15 |
|  | | 8 | 37 | 1.21 | 1.21 | 0.22 |  | 34 | 1.17 | 1.15 | 0.16 |
|  | | 12 | 33 | 1.13 | 1.12 | 0.16 |  | 34 | 1.10 | 1.09 | 0.25 |
| LDL, mmol/L | | |  |  |  |  |  |  |  |  |  |
|  | | 0 | 39 | 2.39 | 2.40 | 0.44 |  | 37 | 2.10 | 2.04 | 0.36 |
|  | | 4 | 37 | 2.49 | 2.31 | 0.45 |  | 35 | 2.09 | 2.10 | 0.41 |
|  | | 8 | 39 | 2.69 | 2.68 | 0.54 |  | 34 | 2.39 | 2.37 | 0.51 |
|  | | 12 | 33 | 2.81 | 2.95 | 0.50 |  | 34 | 2.57 | 2.62 | 0.60 |

Abbreviations: HDL, high-density lipoprotein; LDL, low-density lipoprotein

**Table S3.** Cox Regression Model Estimates of Risk for Time to Appetite Increase by Treatment Groups

|  |  |  | DF | Beta | SE | Chi-Sq | P | Hazard ratio |
| --- | --- | --- | --- | --- | --- | --- | --- | --- |
|  | Gender | Male | 1 | -0.26 | 0.37 | 0.501 | 0.4790 | 0.770 |
|  | Age (years) |  | 1 | -0.092 | 0.041 | 5.033 | 0.0249 | 0.912 |
|  | Duration of illness (month) |  | 1 | -0.021 | 0.048 | 0.181 | 0.6704 | 0.980 |
|  | olanzapine |  | 1 | -0.017 | 0.102 | 0.029 | 0.8642 | 0.983 |
|  | Treatment group | Olan+ | 1 | -0.53 | 0.34 | 2.403 | 0.1211 | 0.589 |

Abbreviations: Olan+, olanzapine plus probiotics

**Table S4**. Antipsychotic Treatment Effect on Psychopathological Symptoms

|  | |  | Baseline | |  | After treatment | |  | Group difference | | | |  |
| --- | --- | --- | --- | --- | --- | --- | --- | --- | --- | --- | --- | --- | --- |
|  | |  | LS mean | SE |  | LS mean | SE |  | Beta | SE | 95%CI | *P* | *P*^a^ |
| **PANSS total** | | |  |  |  |  |  |  |  |  |  |  |  |
|  | Olanzapine + | | 80.18 | 1.29 |  | 55.67 | 1.33 |  | 24.51 | 1.79 | 21.0, 28.0 | <.0001 | 0.48 |
|  | Olanzapine | | 79.50 | 1.31 |  | 56.8 | 1.31 |  | 22.7 | 1.82 | 19.1, 26.3 | <.0001 |  |
| **Positive** | | |  |  |  |  |  |  |  |  |  |  |  |
|  | Olanzapine+ | | 28.76 | 1.05 |  | 14.67 | 1.08 |  | 14.09 | 1.46 | 11.2, 17.0 | <.0001 | 0.64 |
|  | Olanzapine | | 28.75 | 1.07 |  | 13.7 | 1.07 |  | 15.05 | 1.48 | 12.1, 18.0 | <.0001 |  |
| **Negative** | | |  |  |  |  |  |  |  |  |  |  |  |
|  | Olanzapine+ | | 21.93 | 0.95 |  | 17.56 | 0.98 |  | 4.37 | 1.32 | 1.8, 7.0 | 0.0012 | 0.61 |
|  | Olanzapine | | 23.37 | 0.97 |  | 19.97 | 0.97 |  | 3.41 | 1.34 | 0.8, 6.0 | 0.012 |  |
| **General** | | |  |  |  |  |  |  |  |  |  |  |  |
|  | Olanzapine+ | | 29.49 | 1.05 |  | 23.44 | 1.08 |  | 6.05 | 1.46 | 3.2, 8.9 | <.0001 | 0.39 |
|  | Olanzapine | | 27.37 | 1.07 |  | 23.13 | 1.07 |  | 4.24 | 1.48 | 1.3, 7.1 | 0.0048 |  |

Abbreviations: Olanzapine+, Olanzapine plus probiotics.

LS mean, post-hoc least square estimate of mean; P^a^, P-value for testing the difference in the treatment effect between two treatments.

**Table S5.** Estimates of Treatment Effect on Lipid Profiles Over Time by Treatment Groups

|  | Olanzapine+ | |  | Olanzapine | |  | Group difference | | | |  |
| --- | --- | --- | --- | --- | --- | --- | --- | --- | --- | --- | --- |
|  | LS Mean | SE | *P* | LS Mean | SE | *P* | Beta | SE | 95%CI | *P* | *P*^a^ |
| TG, mmol/L |  |  | <.0001 |  |  | <.0001 |  |  |  |  | 0.208 |
| Baseline | 0.83 | 0.067 |  | 0.72 | 0.068 |  | 0.112 | 0.094 | -0.072,0.296 | 0.2372 |  |
| 4 weeks | 0.86 | 0.068 |  | 0.87 | 0.069 |  | -0.004 | 0.095 | -0.190,0.182 | 0.9663 |  |
| 8 weeks | 1.08 | 0.068 |  | 1.08 | 0.069 |  | -0.007 | 0.095 | -0.193,0.179 | 0.9457 |  |
| 12 weeks | 1.26 | 0.069 |  | 1.26 | 0.069 |  | -0.001 | 0.096 | -0.189,0.187 | 0.9884 |  |
| Chol, mmol/L |  |  | 0.0009 |  |  | <.0001 |  |  |  |  | 0.028 |
| Baseline | 3.69 | 0.099 |  | 3.67 | 0.1 |  | 0.026 | 0.138 | -0.244,0.296 | 0.8504 |  |
| 4 weeks | 3.79 | 0.1 |  | 3.53 | 0.101 |  | 0.263 | 0.139 | -0.009,0.535 | 0.0614 |  |
| 8 weeks | 3.91 | 0.1 |  | 3.8 | 0.102 |  | 0.104 | 0.14 | -0.170,0.378 | 0.4571 |  |
| 12 weeks | 3.99 | 0.102 |  | 4.04 | 0.102 |  | -0.054 | 0.141 | -0.330,0.222 | 0.7037 |  |
| HDL, mmol/L |  |  | 0.0003 |  |  | <.0001 |  |  |  |  | 0.6 |
| Baseline | 1.22 | 0.031 |  | 1.23 | 0.032 |  | -0.011 | 0.044 | -0.097,0.075 | 0.7928 |  |
| 4 weeks | 1.25 | 0.032 |  | 1.23 | 0.032 |  | 0.025 | 0.044 | -0.061,0.111 | 0.5728 |  |
| 8 weeks | 1.22 | 0.032 |  | 1.17 | 0.033 |  | 0.049 | 0.045 | -0.039,0.137 | 0.2789 |  |
| 12 weeks | 1.16 | 0.033 |  | 1.09 | 0.033 |  | 0.023 | 0.045 | -0.065,0.111 | 0.6095 |  |
| LDL mmol/L, |  |  | <.0001 |  |  | <.0001 |  |  |  |  | 0.55 |
| Baseline | 2.38 | 0.081 |  | 2.09 | 0.083 |  | 0.285 | 0.114 | 0.062,0.508 | 0.0136 |  |
| 4 weeks | 2.47 | 0.083 |  | 2.08 | 0.084 |  | 0.395 | 0.116 | 0.168,0.622 | 0.0009 |  |
| 8 weeks | 2.68 | 0.081 |  | 2.37 | 0.085 |  | 0.313 | 0.116 | 0.086,0.540 | 0.0077 |  |
| 12 weeks | 2.78 | 0.085 |  | 2.54 | 0.085 |  | 0.237 | 0.118 | 0.006,0.468 | 0.0471 |  |

Abbreviations: TG, triglycerides; Chol, total cholesterol; HDL, high-density lipoprotein; LDL, low-density lipoprotein;

LS mean, post-hoc least square estimate of mean;

*P*^a^, *P* value for testing the difference in lipid profiles over time between treatments

.

**Table S6.** Treatment Effect on Antipsychotics-induced Metabolic Disturbance after Adjusting for Appetite Increase

|  | | Olanzapine+ | |  | Olanzapine | |  | Group difference | | | |  |
| --- | --- | --- | --- | --- | --- | --- | --- | --- | --- | --- | --- | --- |
|  | | LS Mean | SE |  | LS Mean | SE |  | Beta | SE | 95%CI | *P* | *P*^a^ |
| **Fasting insulin, uIU/mL** | | |  |  |  |  |  |  |  |  |  |  |
| Baseline | 7.85 | | 0.55 |  | 7.72 | 0.53 |  | 0.14 | 0.75 | -1.33, 1.61 | 0.858 | <.0001 |
| 4 Weeks | 7.66 | | 0.55 |  | 8.23 | 0.54 |  | -0.58 | 0.76 | -2.07, 0.91 | 0.443 |  |
| 8 Weeks | 8.26 | | 0.56 |  | 10.21 | 0.54 |  | -1.94 | 0.76 | -3.43, -0.45 | 0.012 |  |
| 12 Weeks | 10.56 | | 0.56 |  | 13.64 | 0.54 |  | -3.07 | 0.76 | -4.56, -1.58 | <.0001 |  |
| **HOMA-insulin resistance** |  | |  |  |  |  |  |  |  |  |  |  |
| Baseline | 1.55 | | 0.12 |  | 1.51 | 0.11 |  | 0.035 | 0.16 | -0.28, 0.35 | 0.826 | 0.0012 |
| 4 Weeks | 1.48 | | 0.12 |  | 1.63 | 0.12 |  | -0.15 | 0.16 | -0.46, 0.16 | 0.352 |  |
| 8 Weeks | 1.63 | | 0.12 |  | 2.05 | 0.12 |  | -0.43 | 0.16 | -0.74, -0.12 | 0.009 |  |
| 12 Weeks | 2.24 | | 0.12 |  | 2.77 | 0.12 |  | -0.53 | 0.16 | -0.84, -0.22 | 0.002 |  |
| **Body mass index, kg/m^2^** |  | |  |  |  |  |  |  |  |  |  |  |
| Baseline | 21.44 | | 0.35 |  | 21.54 | 0.35 |  | -0.1 | 0.49 | -1.06, 0.86 | 0.84 | 0.143 |
| 4 Weeks | 21.94 | | 0.35 |  | 22.42 | 0.35 |  | -0.48 | 0.49 | -1.44, 0.48 | 0.327 |  |
| 8 Weeks | 23.4 | | 0.35 |  | 23.92 | 0.35 |  | -0.52 | 0.49 | -1.48, 0.44 | 0.294 |  |
| 12 Weeks | 24.41 | | 0.36 |  | 24.54 | 0.35 |  | -0.12 | 0.49 | -1.08, 0.84 | 0.806 |  |
| **Weight, kg** |  | |  |  |  |  |  |  |  |  |  |  |
| Baseline | 57.58 | | 0.89 |  | 59.01 | 0.89 |  | -1.44 | 1.24 | -3.87, 0.99 | 0.249 | 0.11 |
| 4 Weeks | 58.86 | | 0.89 |  | 61.36 | 0.89 |  | -2.51 | 1.24 | -4.94, -0.08 | 0.046 |  |
| 8 Weeks | 62.6 | | 0.9 |  | 65.2 | 0.9 |  | -2.61 | 1.24 | -5.04, -0.18 | 0.039 |  |
| 12 Weeks | 65.19 | | 0.9 |  | 66.8 | 0.9 |  | -1.61 | 1.25 | -4.06, 0.84 | 0.201 |  |

*P*^a^, the *P*-value for testing the overall treatment effect by the status of appetite increase;

LS Mean, post-hoc least square estimate of mean; SE, standard error.

**Figure S1.** Probability of time to appetite increase between two-treatments (group=1, olanzapine plus probiotics; group=2, Olanzapine only)

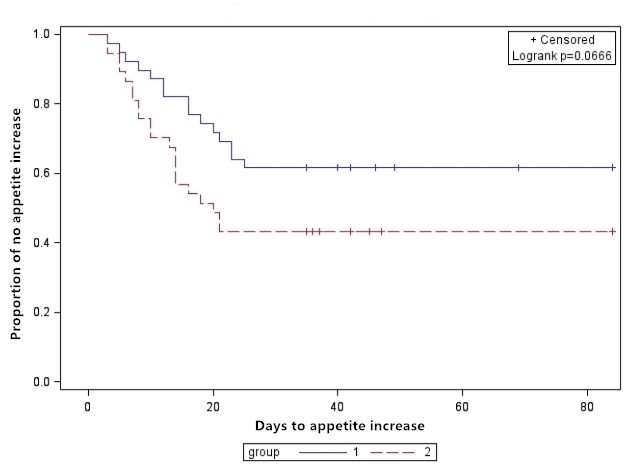
